# Patient Journey: A Qualitative Study of Hepatocellular Carcinoma Patient Experience

**DOI:** 10.1101/2025.03.04.25320693

**Authors:** Michelle Liu, Matthew Loxton

## Abstract

Patients living with hepatocellular carcinoma (HCC) face significant barriers to receiving quality care, including delays in diagnosis and treatment, disease stigma, and financial challenges. This paper develops a three-part measurement system to categorize patient experiences of the care journey. Patient experiences were classified in terms of grounded constructs such as education, self-advocacy, and medical record utilization, as well as integrated care teams, including support groups, counselors, dietitians, palliative care providers, and navigators. These constructs were additionally cross-categorized in terms of care quality and root cause.

## Method

While many care-quality studies have been published, and many delivery metrics are in use, few have focused on the patient and caregiver perspectives, and even fewer metrics are derived directly from those perspectives. The objective of this study is to develop an initial taxonomy of care journey experiences as seen from the patient and caregiver point of view and to give voice to those patients and caregivers who participated. This qualitative study used semi-structured interviews that were conducted virtually through Microsoft Teams and is not intended to be generalized to the entire population of HCC patients, nor to the general population of people living with cancer.

Since generalization was not a goal of this study, we used a combination of convenience and snowball sampling. We used existing HCC patient-advocate contacts and forums to announce the study and invite participation, and repeated this announcement on social media. Since some participants were anticipated to be of low socioeconomic status, an honorarium of $100 was provided in return for an interview of up to 90 minutes. The recruitment goal was n=7-15 participants, and we closed recruitment once we had a commitment from 14 participants.

A standardized greeting was read to the participant at the start of each session, which stated the research purpose, and identified the two research team members and their roles. We explained participant rights, including the right to leave the project at any time, and we requested permission to record the session for research purposes. We explained that these recordings would be deleted at the conclusion of the study and that their identities would not be revealed in any reports or papers. For record-keeping purposes, permission to record was again confirmed after the recording was started. Only one participant declined to be recorded but agreed to have a transcript generated. Transcripts were generated automatically by the Microsoft Teams AI transcription feature and were manually edited for accuracy and validity. Transcripts and audio recordings were imported into a qualitative data analysis tool.^1^

Both researchers independently read the transcripts and used the linked timestamped audiovisual files to verify or clarify understanding, before adding memos to describe the context and implications of text segments and capture any thoughts on potential coding. We subsequently reviewed all memos and developed grounded codes that categorized and captured the key points made by participants. We held a brainstorming session to refine coding, using a card-sorting methodology to form parent codes and code families. After the family of grounded codes was established, these codes were used in collaborative cycles, using MAXQDA TeamCloud. During the seven collaborative cycles, some codes were merged or refined, but the intention was to create a broad code taxonomy to represent the greatest number of insights. The MAXQDA AI tool was used to develop a summary for all the coded segments for each code. This was used to test if our code scope was complete, and the summary was added to the codebook.

After the analysis in MAXQDA was complete, we exported the coded segments, AI summaries, and memos to MS Word to develop a report.

## Discussion

Four grounded categories (Figure 1) emerged from participant descriptions of the care journey and its features. Each of these parent categories contained subcategories that further explored the facets of what participants communicated (number of sub-categories given in parentheses)

**Figure 1.**
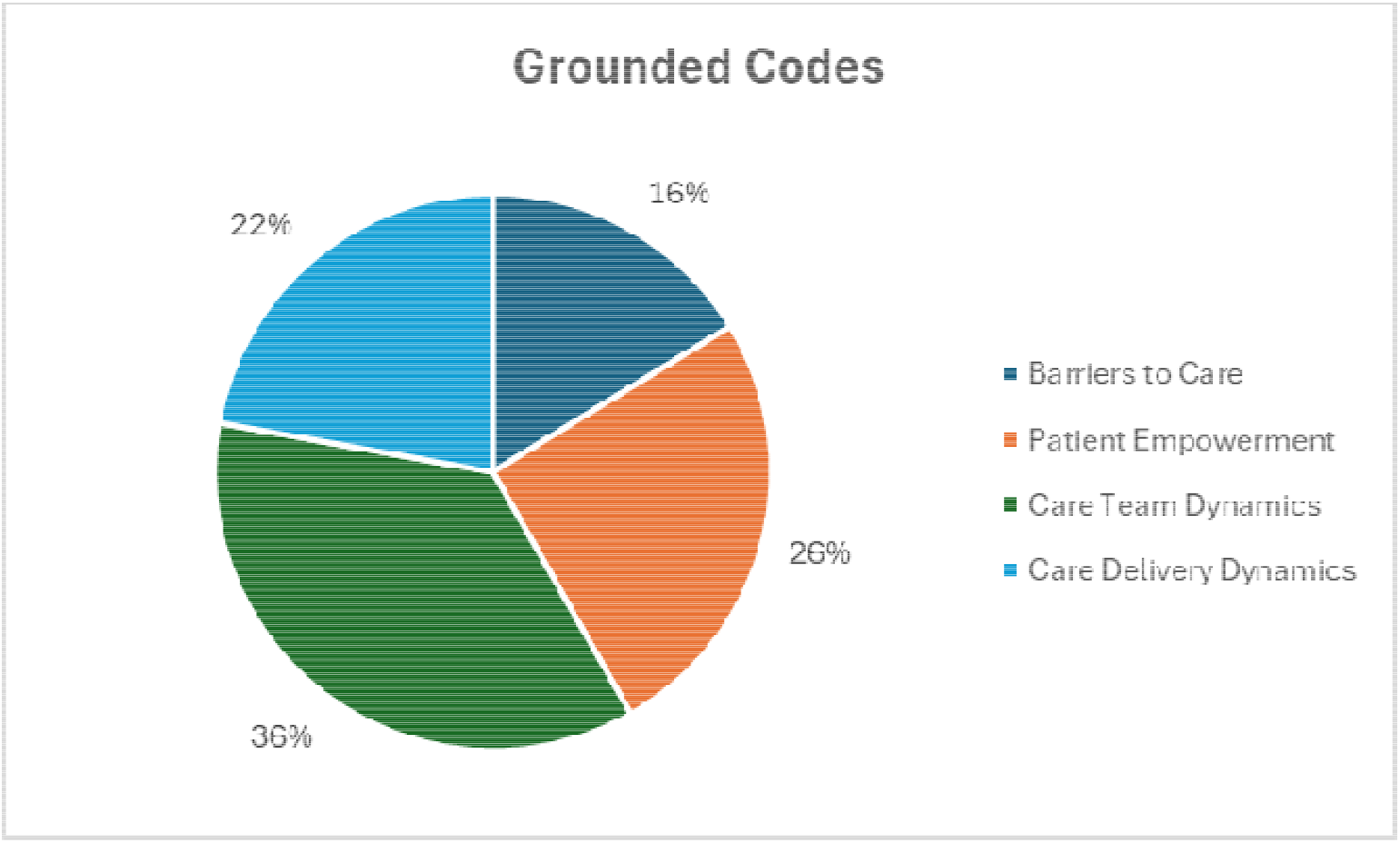
Grounded Code Categories.

1. Barriers to Care (5)
2. Patient Empowerment (6)
3. Care Team Dynamics (7)
4. Care Delivery Dynamics (5)

An “effect family” of codes (Figure 2) was created by adding “accessibility” as a construct to the Institute of Medicine (IOM) “domains of health care quality” (IOM, 2001)). This allowed us to link each care outcome quality dimension to the grounded codes, and address the question of what benefit or harm is represented in the care journey. Tying a standardized care quality dimension to each grounded segment may allow other researchers in that field to better use our findings, and to allow this research to be used to better understand care delivery from a patient’s view of quality that will map to industry standards. The category that attracted the highest number of coded segments was “Patient Centered” (29%), and combined with “Effective” (25%) and “Accessible” (17%), accounted for 71% of coded segments across all participants.

**Figure 2.**
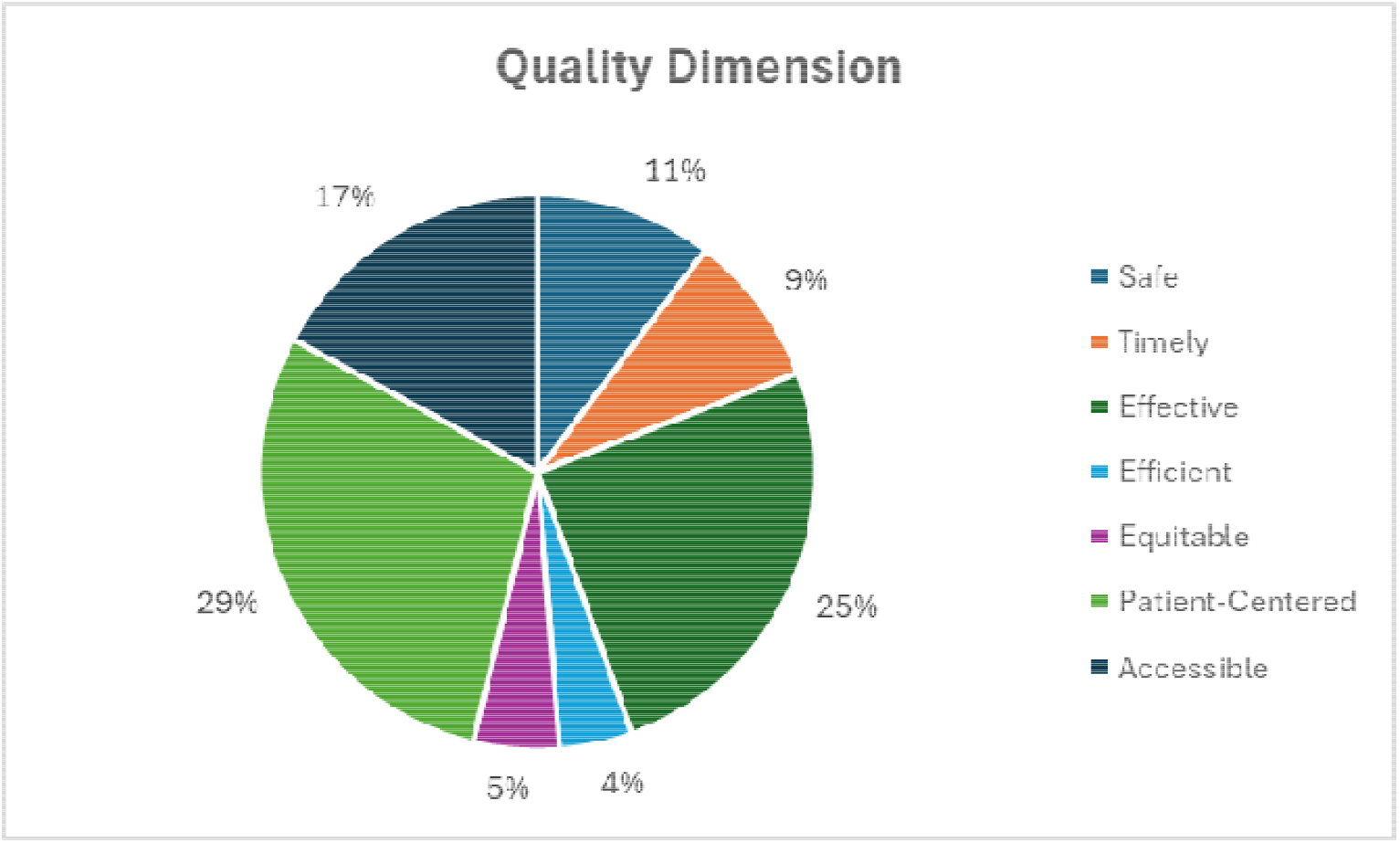
Quality Dimensions Categories.

To provide an element of causation, we mapped coded segments to a pre-existing set of Lean Six Sigma “root cause” dimensions (Figure 3). This approach was taken to make our research more usable to quality improvement groups in healthcare, and to provide a much-needed patient perspective of care quality. The category of “People” (52%) as a root cause made up the majority of coded segments, and combined with “Methods & Policy” (28%) and “Materials” (12%), made up 92% of the root causes across all participants.

**Figure 3.**
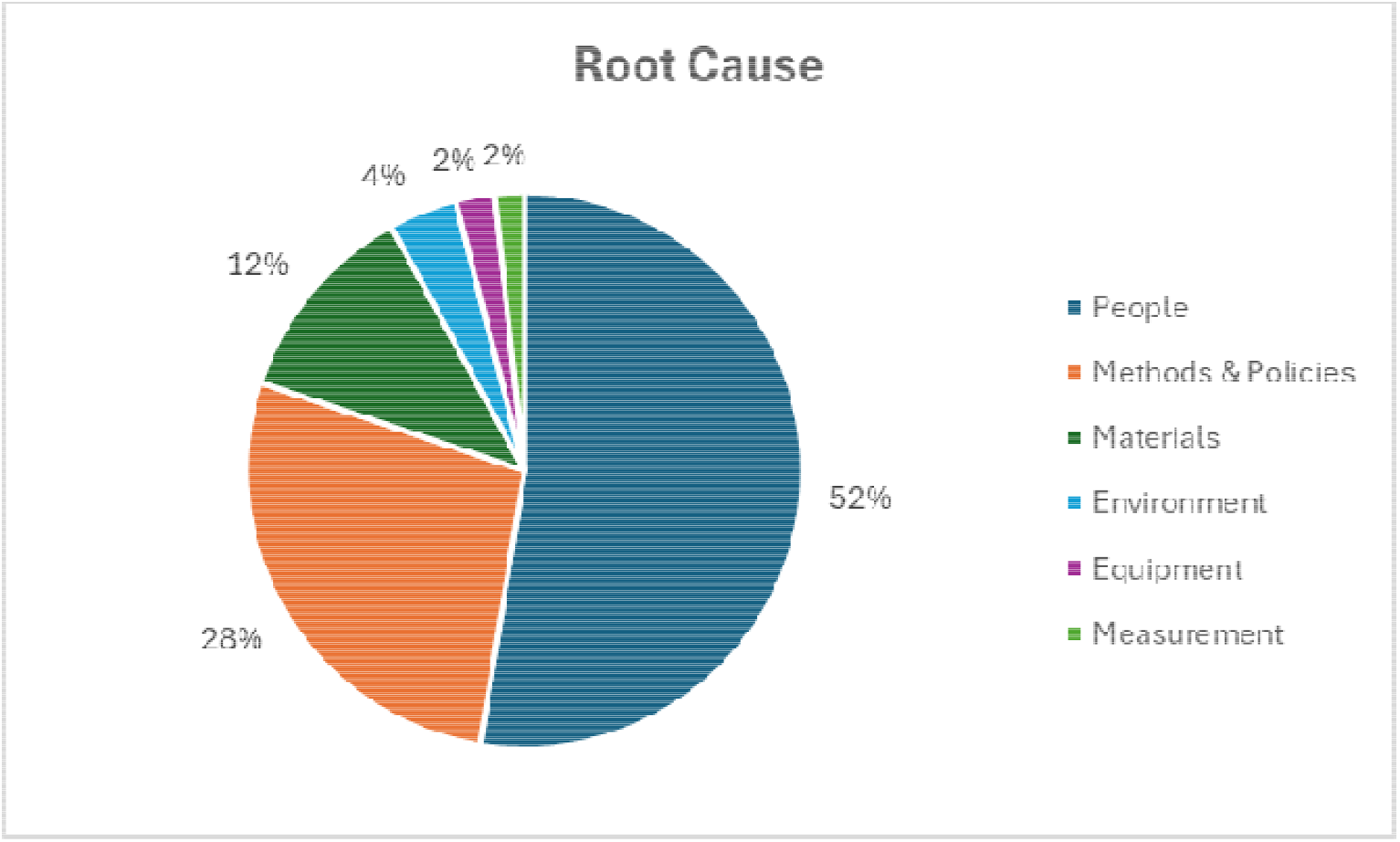
Root Cause Dimensions Categories.

### 2.1. Barriers to Care

Under the category of “Barriers to Care” for patients with HCC, we included any policies, processes, or events that delayed diagnosis, created difficulty in accessing specialists or services, insurance issues, misdiagnosis, and side effects from treatment, but also financial and emotional challenges faced by patients and caregivers. Participants described the effects on their experience of care delays and missed opportunities, the side effects of the treatment, the stigma attached to HCC, the risk of misdiagnosis, and the role that money and funding played in their care journey. From the participants, we discovered that healthcare methods and policies, such as insurance rules, transplant list management, and availability of information were the most predominant causes of care delays and barriers to care timeliness and accessibility. In Figure 4, a heat map chart shows the number of times one of the root causes or quality dimensions intersected with one of the grounded themes that emerged from participant descriptions of their care journey. For example, Methods & Policies and Care Delays coincided 23 times and is highlighted in red for being significantly higher than other intersections.

**Figure 4.**
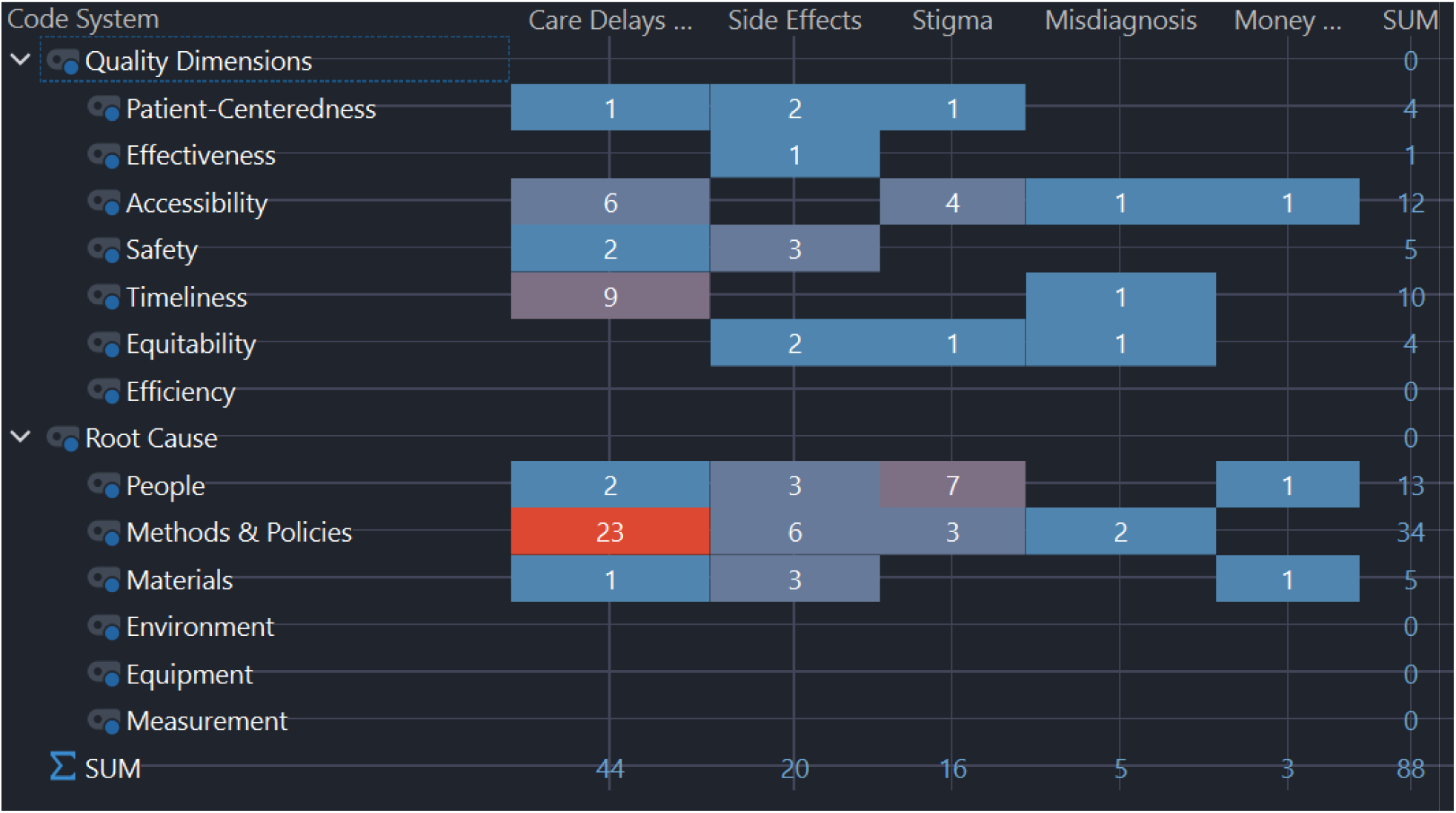
Barriers to Care.

Participants described emotional and physical challenges, such as anxiety, sensitivity to medication, and managing other health conditions. They described hurdles resulting from a lack of information and poor communication from healthcare providers and between providers. Many participants reported that they found a need to advocate for themselves and to do their own research to understand their treatment options and the practical implications of their treatment and condition. In addition, they highlighted the financial and emotional challenges for both patients and caregivers, such as the need to balance caregiving responsibilities with work, and the impact of illness on mental health. The stigma surrounding liver cancer was also described as a barrier to care quality. Finally, the participants discussed the side effects of treatments, such as radiation and immunosuppression and the effect on the care journey.

### 2.2. Patient Empowerment

Patient empowerment is a critical component of informed clinical decision-making and increases patient ability to be treatment adherent (Aslani, 2013), and in healthcare system reform (Segal, 1998). Participants described the influence on their care journey played by patient education, staying physically and mentally active, the degree of self-advocacy and self-motivation, and the ability to ask questions and maintain their own medical records. The World Health Organization (WHO) describes patient empowerment as both an individual and a community or societal process with four core components: (a) Patient understanding their role, (b) Patient having sufficient knowledge to engage meaningfully with their provider, (c) Patient having skills to engage with the healthcare system and attend to their own self-care tasks, and (d) an environment that facilitates access to care that is safe, timely, effective, efficient, equitable, and patient-centered. (WHO, 2009). The participants explained that in practice, these principles were not typically embodied in the care they received. Figure 5 illustrates that participants described issues predominantly related to People, Methods & Policies, and Materials related to Patient Education. Secondarily, they spoke of People factors related to Staying Active and Self Advocacy.

**Figure 5.**
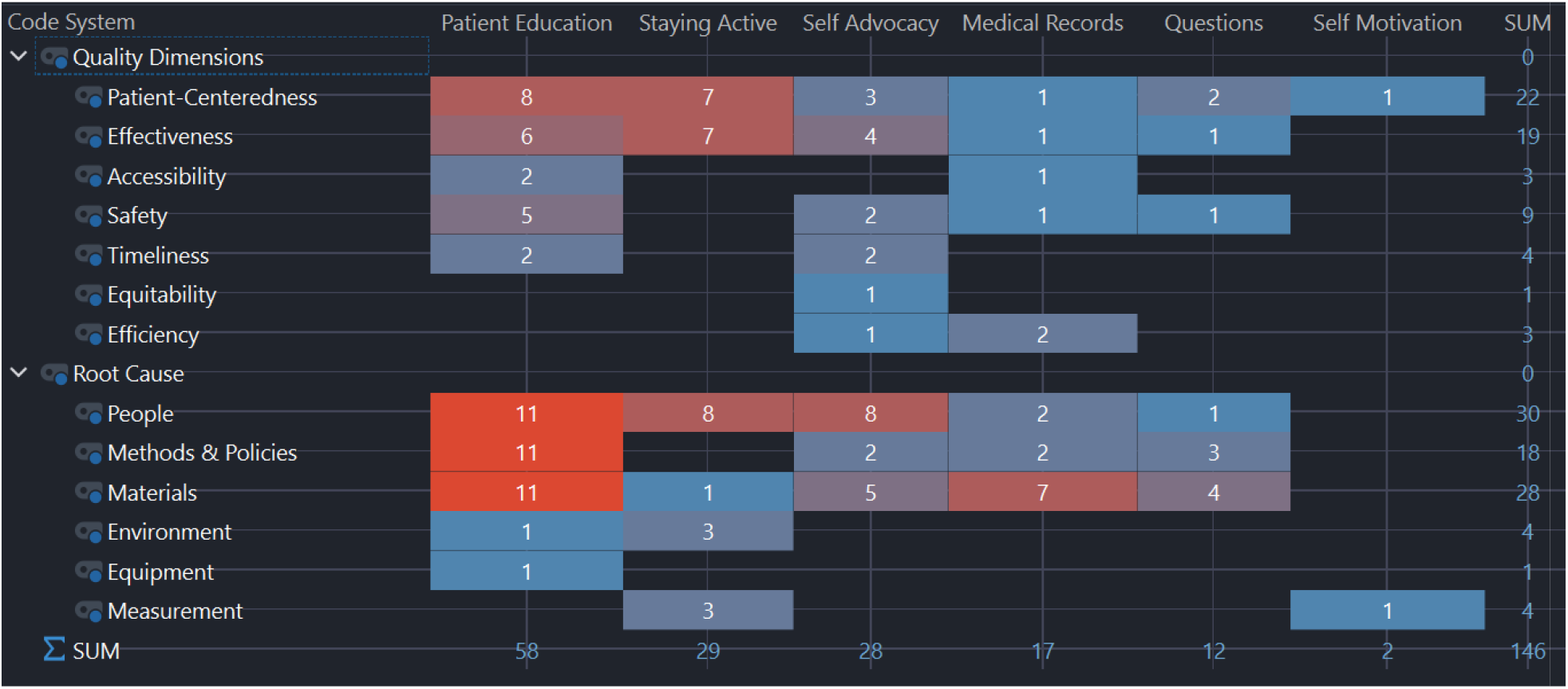
Patient Empowerment.

Participants described an unmet need for trustworthy and actionable information related to the diagnosis, treatment, side effects, self-care needs, and effects on work and relationships of their care and the cancer they are living with. They reported experiencing knowledge gaps pertinent to adhering to care plans, navigating the healthcare environment, and having the knowledge and insights needed to address self-care, risks, and opportunities.

### 2.3. Care Team

A care team comprises the core group of clinicians, technicians, and support people, as well as caregivers, and may vary in composition from patient to patient. Participants outlined the benefits and risks of counseling and therapy, the need for specific oncology dietary support, the need for early discussion and planning for palliative care, and the benefits of access to a medical navigator. Figure 6 illustrates that people, methods & policies, and materials featured strongly in participant explanations, and were associated with outcomes related to patient-centeredness, effectiveness, accessibility, and patient safety.

**Figure 6.**
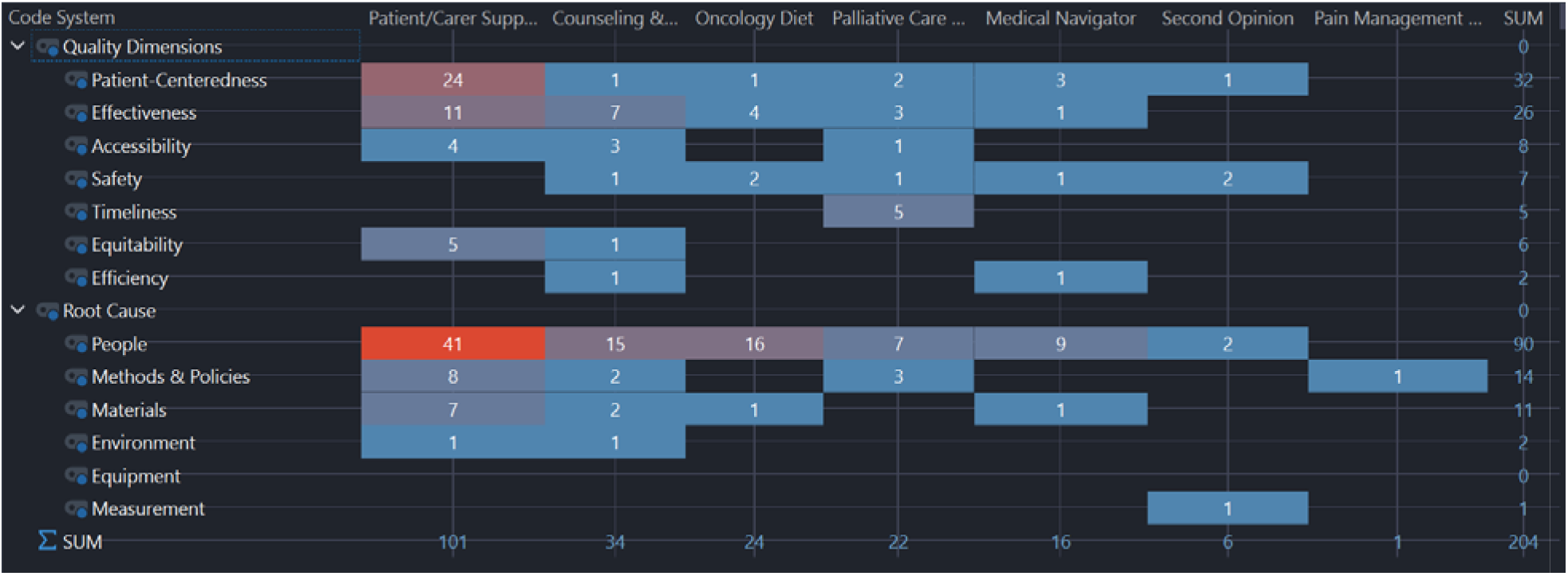
Care Team.

Participants reported belonging to liver cancer and therapy-specific support groups that provided practical information, emotional support, a sense of belonging, and realistic information about the practicalities of the care journey. They reported benefits from these groups, including peer-to-peer support and cancer-specific counselors when faced with persistent anxiety, and episodes of depression resulting from the stress of an HCC diagnosis. Participants described finding great benefits when they had access to a knowledgeable medical navigator to guide them through the local facility policies and programs. They explained the need to seek a second opinion on critical decisions related to their care and prognosis.

### 2.4. Care Delivery

This category relates to participant views on all aspects of the delivery part of care and the implied quality of care. Figure 7 depicts the focus on the people, methods, and policies resulting in mainly patient-centeredness and care effectiveness outcomes. The specific areas of concern relayed by participants included care coordination and teamwork, risk and issue prevention, patient comfort and care ergonomics, and the need for providers to maintain a high index of suspicion to catch HCC at an early stage when symptoms may still be unspecific and diffuse.

**Figure 7.**
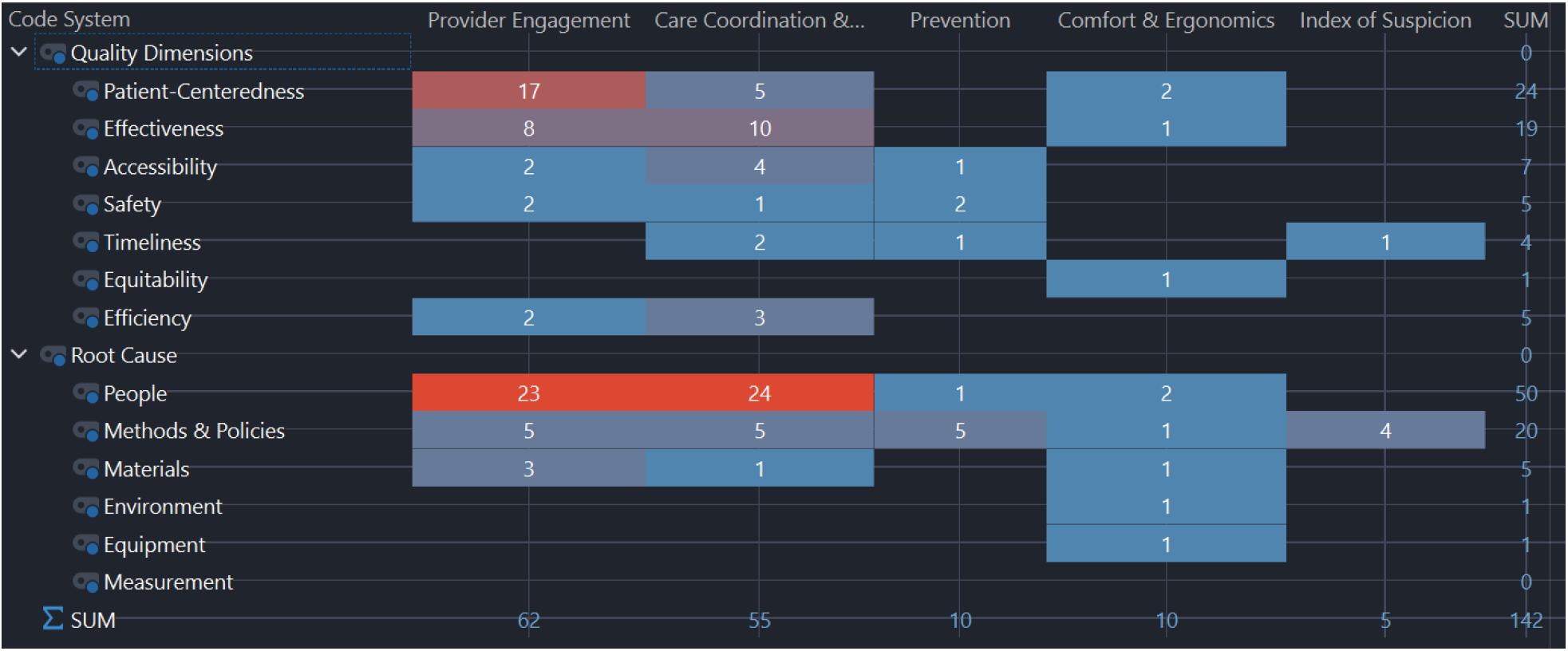
Care Delivery.

Participants described the need for provider engagement and empathy in delivering quality care and highlighted the need for providers to treat them as individuals rather than cases. They outlined the importance of care coordination and integration, and how gaps increased the risk of medical errors. Coordination between medical departments and facilities was an area in which they suggested that the greatest gains could be made and in which they experienced the greatest shortfalls in delivery quality.

## Conclusions

Although many metrics already exist to quantify and guide care delivery, few are derived directly from the patient experience of their care journey. This results in many measurements that guide care delivery and connote care quality in a way that is not representative of the most important stakeholder – the patient. Using qualitative methods to elicit metrics from patients, and overlaying them with industry standards on root cause and quality, may allow quality & safety practitioners to explore ways to improve the patient journey. In this study we developed four families of patient-derived care journey metrics that can be used in this way to improve care for patients living with HCC.

## Data Availability

All data produced in the present study are available upon reasonable request to the authors

## Footnotes

1 MAXQDA version 23, and later migrated to v24.

